# The modified 30-second chair stand test (m-30s-CST) is more sensitive than handgrip strength in detecting muscle strength changes and predicting physical performance and mortality in hospitalized geriatric patients

**DOI:** 10.1101/2025.08.18.25333644

**Authors:** Walther MWH Sipers, Isis Ensink, Martijn JA Rothbauer, Shannon Röhlinger, Audrey HH Merry

**Affiliations:** Department of Geriatric Medicine, Zuyderland Medical Center, Sittard-Geleen, The Netherlands; Department of Physical Therapy, Zuyderland Medical Center, Sittard-Geleen, The Netherlands; Department of Epidemiology, Zuyderland Medical Center, Sittard-Geleen, The Netherlands

**Keywords:** muscle strength, chair stand test, handgrip strength, sarcopenia, physical performance, geriatric patients, mortality

## Abstract

**Objectives:** To compare the modified-30s-Chair-Stand-Test (m-30s-CST) with handgrip strength (HGS) in measuring muscle strength in acutely ill geriatric patients. The aim of this study is to compare the responsiveness and predictive value of the m-30s-CST and HGS for physical performance and two-year mortality in hospitalized geriatric patients

**Methods:** Responsiveness of the m-30s-CST and HGS was assessed in 92 patients (mean age 84±6 y, 53.5% female) by comparing the performance at hospital admission and the day before discharge. These changes were then compared with changes in the ADL-Barthel-Index (ADL-BI) and Short Physical Performance Battery (SPPB).

**Results:** The number of repetitions on the m-30s-CST increased significantly during hospitalization in patients who improved on ADL-BI (n=43) and SPPB (n= 33) and did not change in those who remained stable or worsened (ADL-BI: n= 32 and SPPB: n= 26). There was no significant change in HGS in either patients who improved on respectively ADL-BI (n=43) and SPPB (n=41), nor in those who remained stable or worsened (ADL-BI: n=31 and SPPB: n= 34). Two-year mortality was significantly higher in geriatric patients with low performance on the m-30s-CST. HGS was not associated with 2-year mortality.

**Conclusion:** The m-30s-CST is superior to HGS for assessing changes in muscle strength, serves as a better proxy for physical performance, and is a strong predictor of two-year mortality in hospitalized geriatric patients.

## Introduction

Skeletal muscle mass and strength are strong prognostic factors for the functional decline, morbidity, and mortality of acutely ill hospitalized geriatric patients (1). The European Working Group Sarcopenia Older Peoples-2 (EWGSOP-2) recommends measuring handgrip strength (HGS) or performing the Five times repeated Chair Stand Test (5t-CST) as proxy for muscle strength in diagnosing sarcopenia [8]. In earlier work on this topic, Ensink and coworkers found poor feasibility for the Five Times Sit-to-Stand test (5t-CST), with only 25% of hospitalized geriatric patients able to perform the test. These findings suggest that the 5t-CST is not a feasible tool for evaluating clinical course and treatment during hospitalization. In contrast, they demonstrated good feasibility and excellent test-retest reliability of the Modified Thirty Seconds Chair Stand Test (m-30s-CST) in acutely ill hospitalized geriatric patients. In addition, significant associations were observed between the m-30s-CST and both the Barthel Index for Activity of Daily Living (ADL-BI) and the Short Physical Performance Battery (SPPB), supporting its value as a proxy for functional status in this population” (2). Additionally, they reported a significant association was found between the m-30s-CST and the ADL-BI and the SPPB in a cross-sectional observation. There appeared to be a weak association between HGS and respectively m-30s-CST, ADL-BI and SPPB (2). In geriatric medicine, a muscle strength assessment tool must detect changes during hospitalization to provide insight into physical function and outcome. A a systematic review and meta-analysis from Aarden et al found that HGS decreased during hospital admission (median admission duration 14.7 days) in electively admitted older patients, but not in acutely admitted patients (3). Within that second group (n=343, mean 79±6.6 y, 49% female) they found no significant change in HGS (−0.5kg; P=0.08), but a significant improvement in 5t-CST (+0.7; P<0.001) between admission and 3 months post admission. To the authors’ knowledge, no research has been done on the change in muscle strength and its relation to changes in physical performance and self-reliance during hospitalization in acutely ill geriatric patients. Furthermore, information is lacking on the prognostic value of HGS and m-30s-CST in these physically compromised, frail patients.

In this study, we want to investigate how muscle strength, measured with the m-30s-CST and HGS, changes during hospitalization in patients who improve in ADL-BI and SPPB compared to patients in whom no improvement is found. Additionally, we aim to explore the association between m-30s-CST, HGS and 2-year mortality.

## Materials and methods

### Study sample

All geriatric patients admitted to the acute geriatric ward of the Zuyderland general hospital (the Netherlands) were asked to participate in the study. Recruitment took place over two periods of three months in March 2021 and November 2022. A total of 92 patients were included in the study after admission to the acute care geriatric hospital ward. Inclusion criteria were patients aged above 70 years, a Groningen Frailty Indicator (GFI) score of 4 or greater indicating frailty and having independent mobility (with or without walking aid) before hospital admission [14]. All participants or representatives signed an informed consent form before the start of the study and received written information about the study. Exclusion criteria were being terminal ill with very limited life expectancy, an inability to sit in a standard chair with armrest, not being instructible for performing the tests for any reason or no consent given by patient or representative.

This study complied with the guidelines set out in the Declaration of Helsinki and was approved by the Ethics Committee of Zuyderland and Zuyd Hogeschool, the Netherlands (METCZ20210028).

### Patients’ characteristics

Patient characteristics were retrieved from the medical and nursing files. These included sex, age, diagnosis at hospital admission, medical history, body mass index, nutritional status and frailty score. Carlsson Comorbidity Index (CCI) is a validated tool to predict 10-year survival in patients with multiple comorbidities and is used to categorize the comorbidities (4). The CCI was assessed by a resident of the geriatric department based on the information in the electronic medical file. Bodyweight was measured on a sitting weight scale (SECA, Model 959). Malnutrition was measured using the Short Nutritional Assessment Questionnaire (SNAQ), which is a validated screening instrument for malnutrition. Scores range from 0 to 5; a score of 3 or higher indicates that the patient is malnourished (5). The frailty score was assessed according to the Groningen Frailty Indicator (GFI) criteria, which ranges from 0 to 15: a score of 4 or higher indicates frailty (6).

To study the responsiveness of m-30s-CST and HGS, measurement of these instruments were performed within 3 days after hospitalization and the day before hospital discharge. To study changes in self-reliance and physical performance, respectively the ADL-Barthel Index (ADL-BI) and Short Physical Performance Battery (SPPB) were assessed at these time points. Two-year mortality data were obtained by consulting electronic health records to verify whether patients were alive or deceased, and to document the date of death if applicable. The tests were assessed by the involved researcher, ward resident, physiotherapist or the involved medical student. The tests were carried out in the patients allocated hospital room. Detailed descriptions of the measurement procedures for the m-30s-CST, HGS, ADL-Barthel Index and SPPB can be found in Ensink et al (2025) (2).

### ADL Barthel Index (ADL-BI) and Short Physical Performance Battery (SPPB)

We assume that most patients had already experienced some functional decline due to acute illness before hospital admission. Patients were categorized into two groups; those who improved on the ADL-BI and SPPB and those who deteriorated or remained stable between hospital admission and day before discharge. Given this pre-hospital decline, remaining stable on ADL-BI and SPPB can be interpreted as deterioration.

### Two-year mortality after hospitalization

Information on each patient’s status was retrieved from the hospital electronic medical file to determine whether patients were still alive or were deceased at 2 years after the initial hospital admission, including the exact date of death for survival curve analyses. One researcher retrieved all the information at one time point 2 years after the inclusion of the last patient.

This study was originally powered to test the test-retest reliability. Therefore, we performed a post-hoc analysis to assess the responsiveness of the m-30s-CST compared to changes in the ADL-BI and found a 95.6% level of certainty.

### Statistics

Data analyses were done using IBM SPSS Statistics version 29. Descriptive statistics were used for patient characteristics with means and corresponding standard deviations for continuous variables and percentages for categorical variables. Normal distribution of data (m-30s-CST and HGS) was observed with histograms including skewness and kurtosis and tested using Kolmogorov-Smirnov (KS) test of normality. Repeated Measures ANOVA tests were performed to study changes in m-30s-CST and HGS during hospital stay and in relation with patients’ changes on ADL-BI and SPPB (improvement or stable/worsened).

To test the impact of the m-30s-CST and HGS on the 2-year mortality the Kaplan-Meier Curve was performed. The T-test was used to assess differences between 2-year survivors and non-survivors with respect to: age, BMI, SPPB, m-30s-CST, HGS assessed with Jamar dynamometer, Barthel-ADL score and CCI score. Subsequently, Cox proportional hazard ratio (HR) analysis was performed to determine which of these variables could best predict 2-year mortality. Because of the limited number of patients included and the limited number of ‘events’ (i.e., number of deaths throughout the follow up period), a maximum of 3 covariates were tested at the same time.

## Results

### Study sample

A total of 92 patients were included in the study. Patient recruitment and exclusion with reasons are shown in the flow chart (figure 1).

### Patients’ characteristics

The 92 patients that participated in this study, had a mean age of 84 y ±6SD and a mean GFI score of 6.1± 2.9SD. A total of 49 (53.3%) patients were female.

The baseline patient characteristics, including age, BMI, GFI score, CCI score, SNAQ score, ADL-BI, SPPB, HGS and m-30s-CST were analyzed for differences between patients who improved (n=43) on the ADL-Barthel Index compared to those who remained stable or worsened (n=33) (Table 1). The results indicated that only the CCI score was significantly higher (*P=0*.*009*) in patients who remained stable or worsened on the ADL-BI compared with the patients who improved during hospital stay (Table 1).

### Responsiveness of the m-30s-CST in relation to ADL-Barthel Index

Forty-three patients improved on the ADL-BI during admission. In these patients the number of repetitions on the m-30s-CST increased significantly (*P<0*.*001*) during hospitalization from 3.7± 3.3 to 6.1± 3.2 repetitions. Thirty-three patients worsened or remained stable on the ADL-BI. In these patients the number of repetitions on the m-30s-CST did not change significantly from 4.4± 3.9 to 4.2± 4.1 during hospital stay. Changes in the m-30-s-CST differed significantly (P<0.001) between patients who improved on the ADL-BI and those who worsened or remained stable (Figure 2 Left and Table 2).

### Responsiveness of the HGS in relation to ADL-Barthel Index

Forty-three patients improved on the ADL-BI, showing a non-significant change in HGS during hospitalization from 16.7± 7.8 to 18.4± 8.0 kg. Thirty-three patients worsened or remained stable on the ADL-BI. In these patients the HGS did not change significantly from 18.3± 11.5 to 20.0± 12.8 during hospital stay. Between the patients who improved on the ADL-BI compared with the patients who worsened or remained stable, the changes in the HGS were not significantly different (*P=0*.*068*) (Figure 2 *Right* and Table 2).

### Responsiveness of the m-30s-CST in relation to SPPB

Thirty-three patients improved on the SPPB, showing a significant increase in m-30s-CST repetitions during hospitalization from 3.2± 2.9 to 5.8± 2.5 (*P<0*.*001*). Twenty-six patients worsened or remained stable on the SPPB. In these patients the number of repetitions on the m-30s-CST did not change significantly from 3.8± 3.4 to 4.0± 3.7 during hospital stay. The m-30s-CST difference was highly significant in those who improved on the SPPB versus the patients who worsened or remained stable on the SPPB(P<0.001) (Figure 3 Left and Table 2).

### Responsiveness of the HGS in relation to SPPB

Forty-one patients improved on the SPPB. In these patients the HGS increased non-significantly (*P=0*.*148)* during hospitalization from 18.6± 10.5 to 20.4± 11.9. Thirty-four patients worsened or remained stable on the SPPB, with unchanged HGS during hospitalization (15.5± 8.2 versus 15.3± 6.8). The HGS difference was not significant in those who improved on the SPPB compared with the patients who worsened or remained stable on the SPPB (*P=0*.*069)* (Figure 3 *Right* and Table 2).

### Change in performance during hospital stay

#### ADL-BI and relation with other characteristics

Forty-three patients improved and thirty-three patients remained stable or worsened on the ADL-Barthel Index. There was no significant relation between age, GFI, SNAQ score between the patients who remained stable or worsened compared to those who improved on the ADL-Barthel Index. The patients who remained stable or worsened on the ADL-Barthel Index had a significant higher CCI score (6.7 ± 2.1 versus 5.7 ± 1.7; *P= 0*.*011*) and length of hospital stay (13.8 ± 15.1 versus 6.7 ± 2.1; *P= 0*.*003*) compared to those who improved on the ADL-Barthel Index (Supplementary Table 1).

#### SPPB and relation with other characteristics

Forty-one patients improved and thirty-five patients remained stable or worsened on the SPPB. There was no significant relation between age, CCI, SNAQ score and length of hospital stay between the patients who remained stable or worsened compared to those who improved on the SPPB. The patients who remained stable or worsened on the SPPB had a significant lower GFI score (5.2 ± 2.9 versus 6.7 ± 2.9; *P= 0*.*017*) compared to those who improved on the SPPB (Supplementary Table 2).

#### Muscle strength parameters and 2-year mortality

Two years after hospital admission, forty-nine patients of the 92 patients were deceased. The deceased patients had a significantly lower m-30s-CST at hospital admission (3.5 ± 3.0 versus 5.2 ± 4.4, *P=0*.*014*) compared with those who survived (Supplementary Table 3). Handgrip strength, SPPB, CCI, age GFI, SNAQ and BMI were not significantly different in patients that died within 2 years compared with those patients who survived (Supplementary Table 3).

The Kaplan Meyer survival curves showed significantly higher mortality rates for the patients who were only able to perform three or less compared with the patients who were able to perform 4 or more repetitions on the m-30-CST in hospitalized geriatric patients (Figure 4; Table 3). This findings were similar for the patients who were able to perform 4 or less compared to 5 or more, 5 or less compared to 6 or more repetitions on the m-30s-CST.

There was no significant difference in 2 year survival in patients with low performance on the 5t-CST and handgrip strength compared with patients with a normal performance (Table 3).

#### Cox proportional hazard ratio

Based on the t-test results described above, m-30s-CST, HGS and BMI were included as potential predictors for 2-year mortality in a Cox proportional hazard ratio model. Cox proportional hazard ratio analysis was performed on data for n=92 geriatric patients.

Patients with more repetitions on the m-30s-CST (HR 0.851; CI-95%:0.851-0.996; P=0.040) had a significant lower mortality probability throughout the 2 year follow-up after hospital admission. Patients with higher HGS (HR 0.980; CI-95%: 0.947-1.014; P=0.237) had a non-significantly lower mortality probability throughout the 2 year follow-up after hospital admission.

## Discussion

In this study, we demonstrate that changes in the m-30s-CST – but not in HGS – during hospital stay are significantly associated with changes in physical performance (SPPB) and self-reliance (ADL-BI) in 92 acutely ill hospitalized geriatric patients. Furthermore, we demonstrated that only the m-30s-CST and not HGS was associated with 2 year mortality.

To date, no studies have directly compared the responsiveness of the m-30s-CST and HGS during hospitalization, its associations with changes in SPPB and ADL-BI and predictive value for 2-year mortality. In a cross sectional study located in a rehabilitation center for older adults (n=33), McAllister and coworkers (2020) found a correlation between the m-30s-CST and a modified Barthel-Index with a Spearman rho of 0.737 (P= 0.01). However, they did not study responsiveness of the m-30s-CST. Harris-Love and coworkers found in a cross sectional study that handgrip strength showed a very weak correlation with functional performance (SPPB; ρ=0.260; P=0.012 and ADL-BI; ρ=0.214; P=0.040), without studying the responsiveness of HGS during hospital stay.

In contrary to the findings of Carvalho et al., our present study found no decrease in HGS (18.3± 11.5 versus 20.0± 12.8) during hospitalization in the 31 patients who worsened or remained stable on the ADL-BI (7). They observed a significant decrease in HGS from admission to discharge in hospitalized older surgical patients (n=1168, mean age approximately 70y). These different finding could be explained by differences in patient characteristics and difference in categorizing patients in the present study remaining stable or declining on the ADL-BI versus only patients who declined on the Katz-ADL. Our observation that HGS did not change significantly during hospitalization is in line with the finding by Hartley and co-workers in older acutely hospitalized patients (n=65; median age 84y) (8). They observed a decline in knee-extension strength but not in HGS during hospitalization and a deterioration in general functional ability assessed by means of the ADL-BI from 2 weeks pre-hospitalization and follow-up. Functional mobility, de Morton Mobility Index (DEMMI) improved during hospitalization. However they did not study the relation between knee-extension strength and changes on ADL-Barthel or DEMMI during hospitalization.

Verstraeten and co-workers found that HGS, and not 5t-CST, is useful to predict adverse outcome in geriatric rehabilitation inpatients (9). Contrary to their findings, we observed that HGS is not suitable for predicting mortality. In our study, comparable with the observations done by Verstraeten and co-workers, the majority of patients were unable to perform the 5t-CST (resp 80% versus 76.8%). We believe that the physical capacities of the patients in the study by Verstraeten and co-workers, as well as in our current study, are too limited to perform the 5-times Chair Stand Test (5t-CST): only 23.2% of the 1250 geriatric rehabilitation inpatients (57% female, median age 83 y) were able to perform the 5t-CST and only 56 patients had a normal performance on the 5t-CST. Furthermore, Verstraeten and co-workers concluded that HGS and 5t-CST are not interchangeable as diagnostic measures for suspected sarcopenia. The authors concluded that HGS is suitable for predicting 3 month and 1 year mortality, whereas 5t-CST is less useful for this purpose. In the present study we found that the m-30s-CST, a variant of the CST which overcomes the floor effect of the 5t-CST in these physical compromised patients, is a suitable tool for predicting mortality, unlike HGS. A possible reason for the fact that HGS did not predict mortality is that most of the patients in our study had a handgrip strength below EWGSOP-2 cut-off values (men 22.4 kg SD 8.9 and female 11.3 SD 5.0) and therefore it cannot discriminate for mortality risk. Moreover low performance on the 5t-CST may be partially explained by factors other than sarcopenia alone, such as osteoarthritis, polyneuropathy, and possibly the negative impact of acute inflammation on the neuromuscular junction (10). In our study almost half (n=45) of the patients had documented comorbidities with more or less impact on physical capacity and therefore on the performance on the 5t-CST, such as osteoarthritis (n=45), diabetes (n=27) with probable complications and/or a history of stroke (n=18).

To our knowledge, no studies have investigated the predictive value of the m-30s-CST in terms of mortality. Applebaum and coworkers studied the impact of the m-30s-CST and Timed Up and Go (TUG) on falls in (n=53) older adult Veterans (mean age = 91y, 49 men) residing in a long term care hospital completed in Canada. They found that only the m-30s-CST and not TUG was significantly associated with patients who fall and numbers of falls over 1 year (11).

Improving and retaining self-reliance and overall condition is one of the main goals of acutely ill geriatric hospitalized patients (12). Prioritizing these patient goals underlines the importance of measurements that provide information about changes in physical functioning and not just about mortality risk. The m-30s-CST as a measure of muscle strength in the legs is therefore relevant in acutely ill hospitalized geriatric patients because this test, unlike HGS, shows to be sensitive for changes in physical functioning (SPPB) and self-reliance (ADL-BI) and is a predictor for mortality.

Of the 374 screened patients, only 92 were ultimately included in the study. In view of the high number of patients that are excluded, there may be selection bias. However, we think the probability of this is low as the patient characteristics are similar (data not shown). More than 100 patients had already been discharged early. Reasons for this were diverse, such as no indication for hospitalization, waiting for care, but also a quicker than expected recovery, resulting in discharge to home or another care institution. It may be that we missed patients who had relatively better physical capacities, but also worse. In theory, this could have influenced the impact of changes on the m-30s-CST and changes on the ADL-BI, but we have no data on this. However, we believe this has no significant impact on the association with the 2-year mortality risk. Possible limitations of the study are factors that could have impacted the patient’s ability to perform the m-30s-CST consistently across time: recovery of health throughout the hospitalization, fluctuations, cognitive functioning and physical energy throughout the day and external events that could have impacted the patient’s resilience (13). To avoid this as much as possible, we took the two measurements when we expected the least amount of change in physical and mental conditions. We also tried to take the measurements at the same moment in the patient’s daily schedule so results would not be impacted by fatigue from exercise or mental strain that could have occurred during the day.

We found that completing four or more repetitions on the m-30s-CST was associated with a greater likelihood of survival within two years compared to performing fewer repetitions. However, the exact cut-off value of the number of repetitions on the m-30s-CST as a predictor of mortality needs to be validated in another study.

As the focus of this study was on hospitalized acutely ill geriatric patients, thus concerning frail older patients with high care dependency and low physical performance capacities, caution should be taken when generalizing the findings to other populations. The strength of this study is its conduct in routine hospital practice with acutely ill geriatric patients. In addition, it is the first study that investigates the responsiveness of HGS and m-30s-CST during hospitalization, their relation with changes in self-reliance (ADL-BI) and physical performance (SSPB) and the impact on mortality. In these patients and in this setting we demonstrated that the m-30s-CST, and not HGS, changes during hospital stay in patients who improve on the ADL-BI and SPPB and low performance is related with an increased risk of two year mortality.

In conclusion the m-30s-CST is, unlike HGS, responsive to changes in physical performance (SPPB) and self-reliance (ADL-BI) during hospital stay in the acutely ill hospitalized geriatric population. Furthermore the m-30s-CST is a better indicator for two year mortality compared to HGS. It is an accessible test that quickly gives insight into functional status of hospitalized geriatric patients. More research is needed to access the cut-off values of m-30s-CST in relation to the predict mortality risk.

## Data Availability

All relevant data are within the manuscript and its Supporting Information files.

## Acknowledgements

All authors declare that they have no conflict of interest. No funding was received for conducting this study.

## Ethical standards

All authors comply with the Ethical guidelines for authorship and publishing journal

